# Changes in association between school foods and child and adolescent dietary quality during implementation of the Healthy, Hunger-Free Kids Act of 2010

**DOI:** 10.1101/19005579

**Authors:** Aaron T. Berger, Rachel Widome, Darin J. Erickson, Melissa N. Laska, Lisa J. Harnack

**Author notes:** Corresponding Author: Aaron T. Berger, Division of Epidemiology and Community Health, 1300 South 2nd Street, Suite 300, Minneapolis, Minnesota 55454.

## Abstract

**Purpose:** To estimate the effect of Healthy, Hunger-Free Kids Act of 2010 (HHFKA) implementation on dietary quality of all US school-aged children and adolescents, and examine whether those effects differed by demographic group.

**Methods:** We used survey regression on 2007-2016 National Health and Nutrition Examination Survey (NHANES) data to estimate the proportion of energy intake from school foods and the association between school food intake and dietary quality, before and after HHFKA passage/implementation. To account for demographic changes in the US population over time, inverse probability weighting was employed. The product of the proportion of energy from school foods and the association between school food intake and dietary quality estimated the effect of HHFKA implementation on dietary quality.

**Results:** School food intake quantity remained stable during the study period. HHFKA implementation improved students’ dietary quality by 4.3 Healthy Eating Index-2010 (HEI) points (95% CI: 2.5, 6.1) on days when school foods were eaten, and by 1.3 HEI points (95% CI: 0.73, 1.8) averaged over all days annually.

**Conclusions:** HHFKA implementation improved the total dietary quality of US school students. US students would benefit from eating school meals in the post-HHFKA era, and HHFKA regulations should not be relaxed.

## INTRODUCTION

Poor diet is a leading risk factor related to death and disability in the US.^1^ Though the US is among the wealthiest nations in the world, child and adolescent dietary quality is poor.^2^ Eating a sub-optimal diet has effects that manifest in childhood, such as pediatric obesity^3^ and dental caries.^4^ Additionally, eating habits formed during childhood and adolescence persist into adulthood,^5,6^ affecting lifelong chronic disease risk, and underlining the relevance of childhood diet to health across the life course.

The National School Lunch Program (NSLP) and School Breakfast Program (SBP) were established to support youth nutrition.^7^ The programs have tremendous reach, serving 30 million lunches and 15 million low-cost or free breakfasts each school day.^8^ Students from households with incomes up to 1.85 times the poverty threshold are eligible for meals for free or at a further reduced price.^7^

The Healthy, Hunger-Free Kids Act of 2010 (HHFKA)^9^ was signed into law on December 13, 2010, directing the United States Department of Agriculture (USDA) to reform national standards for all foods served in schools,^10^ better aligning school foods and meal patterns to new dietary guidelines.^11^ Hereafter we will refer to “HHFKA implementation” as the period in which new rules were in effect.

Among other changes, the new rules required schools to offer more servings and greater variety of fruits and vegetables, provide more whole grains, limit fat content in milk, and set calorie and sodium limits.^11^ The standards began July 1, 2012 for school lunches and July 1, 2013 for breakfasts.^11^ Minimum nutritional standards for all other foods sold in schools, including competitive foods (e.g. a la carte items) and snacks, went into effect on July 1, 2014.^12^ Although the Trump administration has rolled back some of these regulations,^13^ each requirement remains more stringent than rules prior to July 1, 2012.

In the six years preceding passage of HHFKA, analyses of National Health and Nutrition Examination Survey (NHANES) data revealed that eating school meals had a beneficial effect on child and adolescent dietary quality, particularly for those with otherwise low-quality diets.^14^ An important question is whether HHKFA successfully optimized this effective program.

Prior evaluations of the dietary effects of HHFKA have examined changes in food selection, consumption, and plate waste in school cafeterias.^15–19^ Encouragingly, students’ cafeteria food selection and consumption shifted to more nutrient-dense, less energy-dense foods,^19^ including more vegetables^15,17,18^ and whole fruits.^15,17,18^ However, one plate waste study in two elementary schools measured reduced fruit and vegetable consumption after HHFKA implementation compared to before.^16^ The USDA’s post-HHFKA report revealed that school meal participants had better dietary quality than matched nonparticipants,^8^ however they made no pre-post HHFKA comparisons.

The goal of this study was to estimate the dietary effects of HHFKA implementation, on days when school foods are eaten, and on an annual, population basis. We also considered whether its effects differed by age and household income.

## METHODS

### Data source and population

Data are from the public use files of the 2007-2016 waves of NHANES, a repeated cross-sectional survey of the civilian, non-institutionalized population of the US.^20^

Participants in this analysis were 4- to 19-year olds who completed two, 24-hour dietary recalls and reported attending K-12 school during the school year (N = 9,532). The final analytic sample was 8,525, as 1,007 participants were missing one or more variables that were needed for sample standardization.

### Dietary assessment

NHANES participants completed questionnaires at home, then visited a mobile examination center (MEC) for a standardized physical examination and in-person 24-hour dietary recall.^20^ NHANES dietary interviews were conducted by proxy (typically a parent) for children up to age 5, and with proxy assistance for ages 6 to 11.^20^ A second dietary recall was completed by telephone on a subsequent day, with 95% occurring 3 to 30 days after the first recall. 24-hour dietary recalls are reliable across telephone and in-person modes^21^ and are considered a valid population-level average of children’s energy intake.^22^

### Outcome: Healthy Eating Index-2010

Healthy Eating Index-2010 (HEI-2010) is a valid and reliable measure^23^ developed to measure adherence to the 2010 Dietary Guidelines for Americans,^24^ the standards that formed the basis for HHFKA regulations.^25^ HEI-2010 includes 12 components. Nine “adequacy components” assess intake of foods where higher intakes are desirable: i.e., vegetables, whole grains, etc. Three “moderation components” assess intake of foods that should be limited: i.e. refined grains, sodium, etc.^23^

We used the per-day scoring algorithm^26^ for each 24-hour dietary recall in the Food Pattern Equivalents Database versions of the NHANES dietary intake files,^27^ to calculate a difference in HEI-2010 score from the first to second 24-hour recall (theoretical range: −100 to 100).

### Exposure: Healthy, Hunger-Free Kids Act of 2010

Schools began to meet HHFKA goals after its passage in 2010, but before legal implementation began in 2012.^28^ Thus this analysis includes three levels of exposure time periods: 1) Pre-passage of the HHFKA (2007-2010 waves), 2) Post-passage, but before implementation of new USDA rules (2011-2012), and 3) Implementation period (2013-2016).

### Exposure: energy from school cafeteria foods

Each food item reported in the 24-hour dietary recall has a caloric value and reported source (e.g., “Cafeteria in a K-12 school”). We calculated the contribution of school foods to total energy intake, summing calories from items reported from “Cafeteria in a K-12 school,” and dividing by total caloric intake from all food items, as done by others.^14^

The exposure was coded as the difference in proportion of energy from school cafeteria foods from the first to the second dietary recall (theoretical range: −100 to 100), allowing participants to effectively serve as their own controls. For example, a student who reported 40% of calories from school meals on Day 1, and 15% of calories from school meals on Day 2, would have an exposure of −25.

### Effect modifiers

Participants who reported eating school meals were asked if the meals were free or reduced-price (FRP), but there was no question on FRP eligibility for those who did not eat school meals. Given this, we used household income at or below 1.85 times the poverty-income ratio as a proxy for FRP eligibility.^7^ For the 9% of participants who did not have a reported income, we substituted self-reported receipt of FRP meals if possible.

“High school-aged” (versus elementary/middle school) was defined as having completed 8th grade or higher at the time of exam, or, if grade level was missing, being age 15 or older.

### Adjustment for confounding by day of week

We assigned a change score of −1 if the first recall occurred on a weekend and the second on a weekday, 1 if the reverse, and 0 if both occurred on weekdays or weekends. A second variable was given a value of 1 if both recalls occurred on weekend days and 0 otherwise. We did not have access to interview dates.

### Demographic and economic factor standardization

Several demographic and economic characteristics, which changed over the observation timeframe, may modify the relationship between HHFKA implementation and dietary quality. We used inverse probability of treatment weights to eliminate the association between time and demographic/economic characteristics, standardizing the five samples to the socio-demographic distribution of the 2015-2016 NHANES cycle.^29–31^

We created treatment weights by fitting a multinomial logistic regression, using the sampling weights provided by NHANES,^31^ to predict the probability that an individual would be observed in each of the five cycles. We used the following predictors: number of persons in the participant’s household (continuous), race/ethnicity (Mexican American/other Hispanic/non-Hispanic white/non-Hispanic black/other race or multiracial), gender (male/female), household reference person’s marital status (currently married/not married), and reference person’s educational attainment (college graduate/lesser attainment). We stratified by two proposed effect modifiers, participant’s age (continuous) and household eligibility for FRP meals (eligible/ineligible).

We created stabilized weights for each NHANES cycle, 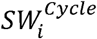, by dividing the predicted probability of being observed in the 2015-2016 cycle, conditional on the modification variables *M* and confounding variables *Z*, by the conditional probability of being observed in the cycle in which the person was actually observed:^29,31^

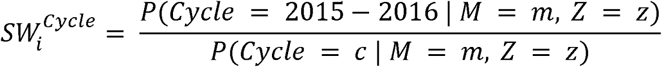

We truncated these weights at the 1st and 99th percentiles. The final analysis weights were the product of the stabilized weights and the NHANES dietary two-day sample weight,^31^ which accounts for demographic/geographic sampling probabilities, non-response to each 24-hour recall, day of the week of the first 24-hour recall, and Day 1 and Day 2 weekday-weekend categories.^32^

### Analysis

School food intake often differed between the two observation days. Many factors that influence school food intake and dietary quality (e.g. family setting, child’s preferences) can be assumed to not change within such a brief period. Therefore, similar to Smith,^14^ we conducted a repeated measures fixed effects analysis assuming that the differences in dietary quality between the two days could be attributed to the substitution of school foods for other foods or vice versa.

First, we estimated the proportion of calories from school foods, both on days when participants reported school foods, and as a proportion of all annual calories. Second, we estimated the effect in each exposure period of each percentage point of calories from school foods. Third, we calculated the scaled estimated effect of food from school on diet quality as: the proportion of energy from school foods times the effect per percentage point. The effect of HHFKA implementation is the change in the effect of school food during the HHFKA Implementation period, compared to the Pre-passage period. We estimated effects overall and for income and grade-level strata.

Both regression steps were conducted in R using the ‘survey’ package^33^ to account for the sampling design and with weighting using the stabilization weights described above. Statistical and mathematical models for each step are included in supplemental materials.

#### Exposure to school foods

After testing for variation by time period, the overall proportion of energy from school foods *E_SF_* was calculated with an intercept-only linear regression:

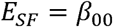

where *β*_00_ is the mean percent of energy from school foods in the total population. The proportions of energy from school foods for each subgroup *G* were estimated with linear regression:

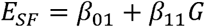

where *β*_01_ is the mean percent of energy from school foods in the reference population (participants not eligible for FRP meals, elementary/middle school students) and *β*_11_ is the difference in energy from school for the subgroup (participants eligible for FRP meals, high school students).

#### Association between school foods and dietary quality

The effect of school food intake on diet quality in the total population, before and after HHFKA implementation, were estimated with weighted linear regression relating the change scores for outcome and exposure by time period:

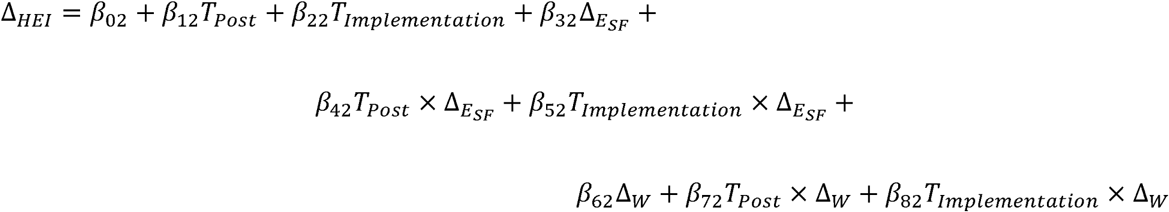

where ∆*_HEI_* is the difference in HEI-2010 score from the first to the second recall, *β*_02_ is the expected difference in HEI-2010 scores between two weekday recalls in the Pre-passage period, if no school food was eaten, and *β*_12_ and *β*_22_ are the changes in that difference in the Post-passage time period *T_Passage_*, and Implementation time period *T_Implementation_*, respectively. *β*_32_ is the difference in HEI-2010 scores for a 1 percentage point increase in energy intake from school cafeteria foods, ∆*_ESF_*, and *β*_42_ and *β*_52_ are the changes in the association between school food intake and HEI-2010 score in the Post-passage or Implementation time periods, respectively. The mean Pre-passage difference in HEI-2010 scores, between weekend and weekday dietary recalls ∆*_W_*, is *β*_62_. The changes in that difference, in the Post-passage and Implementation time periods, respectively, are *β*_72_ and *β*_82_. Because this analysis only uses information from participants whose exposure varies over the two days of dietary recall, we excluded participants with no school food intake in either dietary recall from this analysis step.

For subgroup-specific effects of food from school, we fit the same model, but with the interaction of each term with subgroup *G*.

#### Scaled estimated policy effect

The final step of the analysis was to translate model estimates into individual daily effects, and population-level annual effects. The daily effect is the average effect of school foods for participants who ate school foods, on the day they ate them. Because not every student eats school meals every school day, and school is only in session part of the year, we also estimated a population-level annual effect. This is the average contribution of school foods to the dietary quality of all US students, on all days of the year.

The total estimated effect of the HHFKA on diet is the product of two random variables, estimating first, the proportion of dietary intake from school foods, and second, the change in the effect on HEI score, per percentage point of energy from school foods, in the Implementation period:

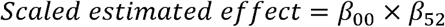

The effect of school meals may also differ by subgroup. For subgroups, the estimated energy intake *β*_01_ or *β*_01_ + *β*_11_ was multiplied by the subgroup-specific change in the effect of food from school on HEI score in the Implementation period. The means and variances for all scaled effects were calculated using standard mathematical formulas for the products and sums of random variables. See supplemental materials for the statistical and mathematical models.

#### Feasibility and sensitivity analyses

Prior to conducting a full analysis of the effect of HHFKA implementation on total dietary quality, we conducted a feasibility analysis to ascertain whether we could detect a difference in nutritional density of school foods in students’ reported lunches. We calculated a Nutrient Rich Foods Index^34,35^ for students’ weekday lunches, in NHANES 2009-2016 dietary recalls. We compared changes in nutrient density of lunches from school cafeterias versus any other source with survey regression.

We conducted three sensitivity analyses. We added a quadratic term to test whether the association between school food intake and dietary quality is linear or exponential, and tested five (versus three) exposure time periods. We compared the fit of the alternative models to those included in this report using nested ANOVA. As an alternative to our unconditional change score model, we also tested an equivalent lagged dependent variable model.^36^

### Statistical reporting

Statistical significance, using a threshold of *P* < 0.05, was used to guide certain modeling choices (e.g. whether average energy intake from school foods differed across cycles), but not to characterize results. All results are presented as means (95% confidence interval [CI]).

## RESULTS

### Participant characteristics

The demographic characteristics of the standardized total sample are shown in Table 1. The average age of participants was 11.5 years. The proportion of non-Hispanic white participants was 48.5%. Almost half of participants were eligible by household income for FRP meals. School foods were reported in at least one recall by 45.7% of participants.

**Table 1.** Descriptive statistics of US school-aged children and adolescents (N = 8,525), NHANES, 2007-2016^a^

|  | mean (SE) |
| --- | --- |
| **Student's characteristics** |  |
| Age (years) | 11.5 (0.1) |
| Gender |  |
| Male (%) | 48.5 (0.9) |
| Female (%) | 51.5 (0.9) |
| Race/Hispanic origin |  |
| Mexican American (%) | 17.3 (1.6) |
| Other Hispanic (%) | 7.7 (0.8) |
| Non-Hispanic White (%) | 48.5 (2.3) |
| Non-Hispanic Black (%) | 15.6 (1.3) |
| Other race (including multi-racial) (%) | 10.9 (0.9) |
| Reported school foods in at least 1 dietary recall (%) | 45.7 (1.8) |
| Daily school cafeteria food intake (kcal) | 185 (8.8) |
| Total daily food intake (kcal) | 1921 (13) |
| HEI-2010 (average of Day 1 and Day 2) | 47.6 (0.24) |
| **Household characteristics** |  |
| Eligible for free/reduced-price meals (%) | 45.9 (1.7) |
| Number of people in household | 4.7 (0.003) |
| **Household reference person's characteristics** |  |
| Age (years) | 41.9 (0.2) |
| College graduate or higher (%) | 28.0 (1.5) |
| Currently married (%) | 68.3 (1.1) |

Abbreviations: HEI-2010, Healthy Eating Index, 2010; kcal, kilocalories; NHANES, National Health and Nutrition Examination Survey.
<sup>a</sup> NHANES cycles standardized to 2015-2016 distribution of participant age, gender, and race/ethnicity, household income-based eligibility for free/reduced-price meals, household size, and household reference person's marital status and educational attainment.

#### Demographic and economic factor standardization

Untrimmed IPTWs had a mean of 1.14 and ranged from 0.28 to 7.81. The mean was greater than 1 due to population growth, between the years being standardized and 2015-2016, the target population. The trimmed mean was 1.13 (range: 0.47 to 2.88). Trimming did not meaningfully change any parameter estimates. Distributions of IPTWs are included in supplementary material.

### Feasibility and sensitivity analyses

The feasibility analysis showed that we could use students’ dietary recalls to detect changes in the quality of school meals. The nutrient density of students’ weekday lunches from school cafeterias increased greatly in the Implementation period, compared to the Pre-passage period. By contrast, the nutrient density of weekday lunches from other sources were virtually unchanged.

The sensitivity analyses did not find improvement of fit using either a nonlinear association between school food intake and dietary quality or a five-level time variable. The alternative, lagged dependent variable specification of the school food-dietary quality regression provided estimates consistent with the unconditional specification at each time point. Feasibility and sensitivity results are included in the supplement.

### Exposure to school foods

Averaged across the full year, including non-school days, 9.9% of all US students’ energy intake came from school cafeteria food (95% CI: 9.0, 10.8) (Table 2, column 8). This is similar to a previous estimate using earlier NHANES cycles.^14^ On days when students reported eating school foods, 33.5% of daily energy was from school foods (95% CI: 32.6, 34.3). Students’ energy intake from school foods did not change statistically significantly across the three time periods.

**Table 2:** Within-Person Association Between Percent of Energy Intake From ’Cafeteria in a K-12 School’ and Change in Healthy Eating Index-2010, Before and After Passage and Implementation of the Healthy, Hunger-Free Kids Act of 2010. NHANES, 2007-2016 (N = 8,525)

|  | Effect of 1 percentage point increase in food energy from school meals on HEI-2010, by time period |  |  | Daily scaled effect of school food and HHFKA implementation on HEI-2010 (school meal participants only) |  |  |  | Annual scaled effect of school food and HHFKA implementation on HEI-2010 (total population, including non-school meal participants) |  |  |  |
| --- | --- | --- | --- | --- | --- | --- | --- | --- | --- | --- | --- |
|  | Pre-Passage (2007-2010) | Post-Passage (2011-2012) | Implementation Period (2013-2016) | Percent of energy intake When school foods are eaten | Daily effect, Pre-Passage | Daily effect, Implementation Period | Daily effect of HHFKA Implementation | Percent of annual energy intake | Total effect, Pre-Passage | Total effect, Implementation Period | Total effect of HHFKA Implementation |
|  | (1) | (2) | (3) | (4) | [(1) × (4)] (5) | [(3) × (4)] (6) | [(6) - (5)] (7) | (8) | [(1) × (8)] (9) | [(3) × (8)] (10) | [(10) - (9)] (11) |
| All school-aged participants | 0.021<br>(-0.015, 0.056) | 0.097<br>(0.039, 0.16) | 0.15<br>(0.11, 0.19) | 33.5<br>(32.6, 34.3) | 0.70<br>(-0.48, 1.9) | 5.0<br>(3.7, 6.1) | 4.3<br>(2.5, 6.1) | 9.9<br>(9.0, 10.8) | 0.21<br>(-0.14, 0.55) | 1.5<br>(1.1, 1.9) | 1.3<br>(0.73, 1.8) |
| By eligibility for free/reduced-price meals (household income ≤ 185% of poverty level): |  |  |  |  |  |  |  |  |  |  |  |
| Not eligible | -0.0019<br>(-0.068, 0.064) | 0.74<br>(-0.0045, 0.15) | 0.15<br>(0.069, 0.23) | 30.3<br>(28.9, 31.7) | -0.059<br>(-2.1, 1.9) | 4.5<br>(2.1, 6.9) | 4.5<br>(1.4, 7.7) | 7.4<br>(6.3, 8.6) | -0.014<br>(-0.51, 0.48) | 1.1<br>(0.48, 1.7) | 1.1<br>(0.32, 1.9) |
| Eligible | 0.036<br>(-0.0015, 0.74) | 0.11<br>(0.018, 0.21) | 0.15<br>(0.12, 0.18) | 36.1<br>(35.1, 37.1) | 1.3<br>(-0.055, 2.7) | 5.5<br>(4.2, 6.7) | 4.2<br>(2.3, 6.0) | 12.8<br>(11.9, 13.7) | 0.46<br>(-0.0021, 0.86) | 1.9<br>(1.5, 2.4) | 1.5<br>(0.81, 2.1) |
| By age group: |  |  |  |  |  |  |  |  |  |  |  |
| Elementary/middle school | 0.039<br>(0.0020, 0.76) | 0.094<br>(0.0090, 0.18) | 0.16<br>(0.11, 0.20) | 33.8<br>(32.8, 34.7) | 1.3<br>(0.067, 2.6) | 5.3<br>(3.6, 6.9) | 3.9<br>(1.9, 6.0) | 11.2<br>(10.2, 12.2) | 0.44<br>(0.020, 0.86) | 1.8<br>(1.2, 2.3) | 1.3<br>(0.62, 2.0) |
| High school | -0.030<br>(-0.13, 0.074) | 0.11<br>(-0.0011, 0.23) | 0.13<br>(0.045, 0.22) | 32.6<br>(30.5, 34.7) | -0.98<br>(-4.4, 2.4) | 4.3<br>(1.4, 7.2) | 5.3<br>(0.86, 9.8) | 7.2<br>(6.2, 8.2) | -0.22<br>(-0.96, 0.53) | 0.95<br>(0.30, 1.6) | 1.2<br>(0.18, 2.2) |
| Abbreviations: HEI-2010, Healthy Eating Index-2010; HHFKA, Healthy, Hunger-Free Kids Act of 2010.<br>All columns present results of survey linear regression or linear combinations of random variables. All analyses adjusted for weekday versus weekend dietary recall date.<br>NHANES cycles standardized to 2015-2016 distribution of participant age, gender, and race/ethnicity, household income eligibility for free/reduced-price meals, household size, and household reference person's marital status and educational attainment |  |  |  |  |  |  |  |  |  |  |  |

Participants eligible for FRP meals consumed more energy from school foods, annually (12.8%, 95% CI: 11.9, 13.7), than did participants who were not eligible (7.4%, 95% CI: 6.3, 8.6). High school students consumed less annual energy from school foods (7.2%, 95% CI: 6.2, 8.2) than younger students (11.2%, 95% CI: 10.2, 12.2).

### Association between school foods and dietary quality

The within-person association between change in school food consumption and change in dietary quality, in all three time periods, is shown in columns 1-3 of Table 2. In the total population in the Pre-passage period, each percentage point of energy from school foods was associated with a 0.021-point increase in the participant’s HEI score (95% CI: −0.014, 0.056). By the HHFKA implementation phase, there was an association of 0.15 HEI points (95% CI: 0.11, 0.19), per percentage point of energy from school foods.

In the Pre-passage period, school foods were beneficial for the diets of students eligible for FRP meals and for elementary and middle school-aged participants, but were not associated with diet quality for those in high school or ineligible for FRP meals. At the Implementation period, each percentage point of intake from school cafeteria foods improved dietary quality by between 0.13 and 0.16 points, for all subgroups. The associations for the whole sample, excluding weekend days, is also depicted in Figure 1. Full model results are included in the supplementary materials.

**Figure 1.**
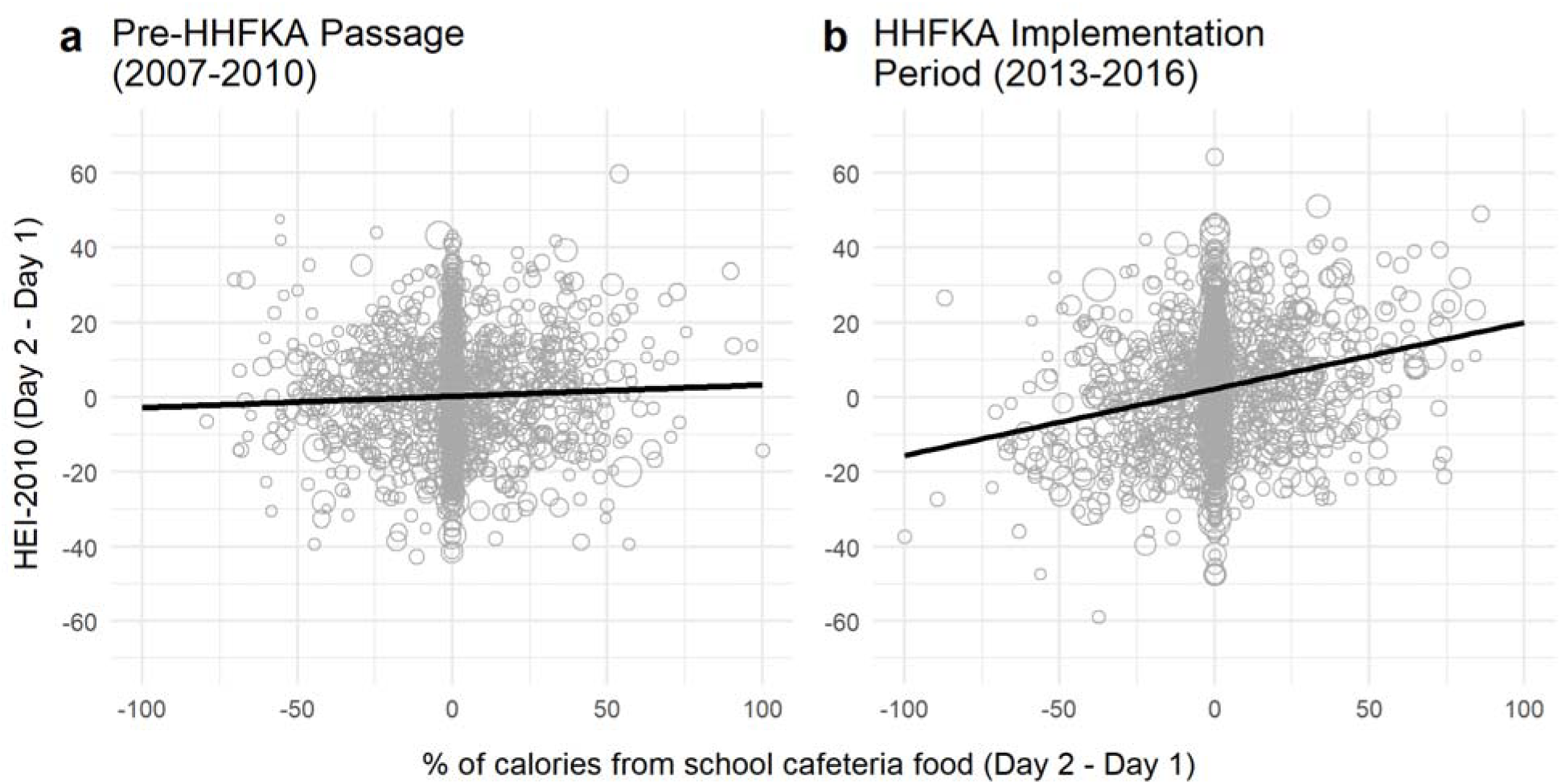
Association between school food intake and dietary quality (HEI-2010) (a) before and (b) during Healthy, Hunger-Free Kids Act of 2010 implementation. Bubble size represents probability weight for participant. NHANES 2007-2010 and 2013-2016, weekend days excluded (N = 2,793)

### Scaled estimated policy effect

Scaled to the relative contribution of school meals to participants’ energy intake, on days school meals were eaten they improved participants’ dietary quality by 0.70 HEI points in the Pre-passage period (95% CI: −0.48, 1.9). By the Implementation period, this increased to 5.0 HEI points (95% CI: 3.7, 6.3). Therefore, HHFKA implementation improved students’ dietary quality by 4.3 HEI points on days when school foods were eaten (95% CI: 2.5, 6.1) (Table 2, column 7).

Scaled to the annual average level of exposure (which includes days school meals were not eaten), implementation of the HHFKA improved the average annual dietary quality of US students by 1.3 HEI points (95% CI: 0.73, 1.8) (Table 2, column 11). The daily and annual scaled improvements in dietary quality attributable to the HHFKA did not differ substantially by subgroup, despite subgroups having different exposure to school foods.

## DISCUSSION

The first four years of HHFKA implementation were associated with substantially improved dietary quality in US students overall and within income and grade-level subgroups. Further, contrary to some predictions that youth would not eat healthier foods, but consistent with other evidence,^19,25^ we found that school food consumption remained stable during HHFKA implementation.

Our findings are concordant with prior assessments of impacts of the HHFKA, that approached the question by analyzing students’ cafeteria food selection and consumption. While analyzing cafeteria food choices demonstrated the healthfulness and desirability of new school foods,^15,17–19^ it could not contextualize policy effects within students’ total diet, which our analysis was able to do. Further, pre-post studies risk unmeasured confounding due to secular trends in youths’ dietary intakes.^2^ This analysis extends the work of Smith,^14^ who had examined how eating school foods was associated with dietary quality prior to the policy change. The fixed effects analysis used in this study protects against that threat to pre-post studies because, at each time point, each participant with varying exposure to school foods served, essentially, as their own “control.”

A limitation of this study is that it does not distinguish between changes in the quality of food from school cafeterias and changes in the quality of food from other sources. Either could increase the apparent effect of school foods on dietary quality. However, the results of the feasibility analysis showed that school lunches improved over time, while lunches from other sources had no changes in nutrient density. This suggests that the cause of the observed improvement was changes in school foods, not foods outside of school. Nevertheless, a causal interpretation of our findings requires the assumption that potential outcomes are independent of HHFKA implementation, given the specified modifiers and confounders.^31^

Inherent in conducting a national observational study without detailed geographic information, we cannot distinguish between effects of federal, state, and local changes in school meal standards. However, the alignment of HHFKA passage and implementation with NHANES cycles points to improvements in school foods being pushed by the policy of interest.

School meals are more beneficial to diet quality in the HHFKA era than they were previously. Communities and families should encourage participation in school meals because they can boost students’ nutrition. Future work could consider what role specific aspects of HHFKA, such as the elimination of unhealthy competitive foods from school cafeterias, plays in dietary quality, in order to further optimize these policies. However, all subgroups we examined will benefit from NSLP/SBP participation, as long as the HHFKA provisions are retained. Federal regulators must resist pressure to roll back HHFKA provisions which would lower school foods’ nutritional value.

## Data Availability

All data used in preparation of this manuscript are publicly available.

https://www.n.cdc.gov/Nchs/Nhanes/

## Acknowledgments

The authors gratefully acknowledge support from the Minnesota Population Center (P2C HD041023) funded through a grant from NICHD.

## Abbreviations

FRP: Free or reduced-price
HEI-2010: Healthy Eating Index - 2010
HHFKA: Healthy, Hunger-Free Kids Act of 2010
NHANES: National Health and Nutrition Examination Survey
NSLP: National School Lunch Program
SBP: School Breakfast Program

